# Frequency of bacterial STI testing amongst people accessing sexual health services in England, 2024: a cross-sectional analysis of national surveillance data

**DOI:** 10.64898/2026.04.08.26349546

**Authors:** George Baldry, Ana-Karina Harb, Lucy Findlater, Dana Ogaz, Stephanie J Migchelsen, Helen Fifer, John Saunders, Hamish Mohammed, Katy Sinka

## Abstract

**Objectives:** There are guidelines in England for recommended sexually transmitted infection (STI) testing frequency, however national estimates for testing frequency are limited. We determined the frequency of STI testing among people accessing sexual health services (SHSs) in England.

**Methods:** We assessed STI testing frequency in face-to-face and online SHSs in England using data from the GUMCAD STI surveillance system. We quantified different combinations of tests within an episode of care and the number of testing episodes completed by each person in 2024 by sociodemographic and behavioural characteristics, as well as clinical setting.

**Results:** Overall, there were 2,222,028 testing episodes at SHSs in 2024 that involved tests for chlamydia, gonorrhoea, syphilis and/or HIV, among 1,750,625 people. Most (66.1%) of these episodes involved tests for all four of these STIs. Most people accessing SHSs in England tested once (80.1%), and a small minority (1.9%) tested at least four times (frequent testers). Frequent testing was more common among gay, bisexual, and other men who have sex with men (GBMSM) (6.7%), London residents (3.6%), online testers (2.5%), people using HIV-PrEP (13%), and people with 5+ partners in the previous 3 months (10.6%). Only 10.5% of GBMSM reporting higher-risk sexual behaviours tested 4+ times despite recommendations for quarterly testing in this group.

**Conclusions:** The majority of those who tested for STIs in England in 2024 only tested once. Frequent testing was more common among GBMSM, people using HIV-PrEP, London residents and people reporting higher risk behaviours. Testing frequency often appears to be aligned with current testing recommendations in England; however, we also observed that a low proportion of GBMSM reporting higher-risk behaviours and HIV-PrEP users tested 4+ times in 2024. It is important to acknowledge groups with lower or higher testing frequency when developing interventions and updating STI testing guidelines.

**WHAT IS ALREADY KNOWN ON THIS TOPIC:** The effectiveness of asymptomatic screening for chlamydia and gonorrhoea in gay, bisexual and other men who have sex with men (GBMSM), and the potential impact of the consequent antibiotic use on antimicrobial resistance has recently been questioned. Testing and treatment remain a key pillar of STI prevention and management; despite this, there is limited evidence of STI testing frequency by people accessing sexual services (SHS) in England.

**WHAT THIS STUDY ADDS:** This analysis shows that the majority of people attending SHSs in England in 2024 tested once, and only a small proportion of people reporting high-risk behaviours tested frequently.

**HOW THIS STUDY MIGHT AFFECT RESEARCH, PRACTICE OR POLICY:** Awareness of groups engaging in high-risk behaviours but are testing infrequently is important to guide interventions and messaging regarding STI testing. The low levels of frequent testing, even among those who would be recommended quarterly testing under UK guidelines, provides important context for wider discussion around asymptomatic STI screening.

## Introduction

Sexually transmitted infections (STIs) represent a major public health concern[1]. In England, asymptomatic HIV and STI screening is recommended to gay, bisexual and other men who have sex with men (GBMSM) at increased risk of STIs[2, 3], and opportunistically to young women or other people with a womb or ovaries through the National Chlamydia Screening Programme (due to the potential adverse reproductive health outcomes[4, 5]). Varying frequencies of asymptomatic testing are recommended for GBMSM based on different criteria[2, 3]. Annual STI testing is recommended for sexually active GBMSM and quarterly testing is recommended for those at higher risk of STI acquisition; including people taking HIV-PrEP, people with over 10 partners in the past year, multiple or anonymous partners since their last STI test, or for people engaging in sexualised drug use[2, 3]. Most STI testing in England involves free face-to-face attendances at sexual health services (SHSs) or online postal self-sampling kits[6, 7].

The rationale for quarterly chlamydia and gonorrhoea screening amongst asymptomatic GBMSM has been questioned[8–11]. Reasons for this include the high proportion of asymptomatic chlamydia and gonorrhoea infections in GBMSM, which are not associated with morbidity and often self-resolve[11]. Also, the evidence that screening reduces the prevalence of chlamydia and gonorrhoea in GBMSM is weak[12]. Increased screening also leads to a higher use of antibiotics, which may contribute to antimicrobial resistance (AMR) without benefitting the individual[10, 12, 13]. This is of great concern given the high levels of AMR in *Neisseria gonorrhoeae, Mycoplasma genitalium* and *Shigella* spp[14, 15]. Despite these concerns regarding frequent testing for chlamydia and gonorrhoea in GBMSM, it is reported that STI testing frequency in GBMSM could be lower than recommended[16, 17]

It is important to gain an understanding of STI testing frequency in everyone attending SHSs in England to accurately inform testing guidelines and aid interpretation of STI epidemiology. Within the context of the UK’s STI testing recommendations, this paper used national surveillance data to describe the frequency of testing in England, and the associated demographic and clinical characteristics of frequent testers.

## Methods

### Data source

This cross-sectional analysis used data from the GUMCAD STI Surveillance System (hereafter ‘GUMCAD’), which includes routinely collected data on SHS attendances (both face-to-face and online) in England. GUMCAD is an electronic, pseudonymised, individual-level dataset which includes demographic and behavioural data on service attendees and has records of all STI tests and diagnoses. In GUMCAD, people have clinic-specific identification codes that allow follow-up over time within, but not between, SHSs. We identified individuals who accessed SHSs in England in 2023 and 2024 and received tests for chlamydia, gonorrhoea, syphilis and/or HIV. We only present data on tests that occurred in 2024; however, tests occurring in 2023 were used to calculate time between tests and to identify any STI diagnoses in the previous year (in the 12 months before the onset of the testing episode). To avoid double-counting (multiple SHS visits for the same infection), only one test per episode (42 days following a test) was considered[18].

Men were coded as GBMSM throughout 2024 if they were recorded as gay or bisexual at any point. Gender identity and sexual orientation categories were defined based on groups identified in current STI testing guidelines[2] (GBMSM, heterosexual men and women [all groups were also stratified based on sexual risk]). We grouped women into a single category due to low numbers of tests in women who exclusively have sex with women. We included data on whether people had symptoms when they were tested (yes/no [there are no data on syndrome/presentation]), and whether they were testing after partner notification as a partner of a diagnosed index case. Where reported, we also included data on the number of sex partners in the 3 months prior to the SHS attendance. Where we have reported unknown values (sexual orientation, number of partners, symptomatic status), this included data where this is not known, not stated and/or not reported. In individual-level analyses, where someone had attended a SHS multiple times but had a different symptomatic status (any combination of symptomatic/asymptomatic/unknown), they have been labelled as having a mixed symptomatic status. We only included episodes that had a test for either chlamydia and/or gonorrhoea in this analysis. HIV status was defined using clinician-coded data for a HIV diagnosis or attending a SHS for HIV-related care (a person was recorded as living with HIV for all episodes following a diagnosis or report of HIV-related care). An individual was recorded as using HIV-PrEP from the point a HIV-PrEP prescription was recorded by a clinician until the point that a stop code was reported or 6 months had passed without a subsequent HIV-PrEP prescription.

### Data analysis

First, we identified different combinations of chlamydia, gonorrhoea, syphilis and HIV tests within an episode. Categories were defined based on clinical coding as: a single test for chlamydia or gonorrhoea; a dual chlamydia and gonorrhoea test; a test for chlamydia, gonorrhoea and syphilis (considered a full sexual health screen for people living with HIV); and full screens (a test for all 4 infections). Any other combination (e.g a chlamydia and syphilis test) were aggregated into a single group. We compared proportions of these tests in face-to-face and online SHSs. The number of online tests were based on online postal self-service kits that were returned by service users.

We defined testing frequency categories based on the number of tests completed by a person in 2024. These categories were 1 test, 2-3 tests and 4+ tests (frequent testing). We examined the demographic, behavioural and clinical characteristics of individuals testing in 2024 by testing frequency category. We also calculated the median and interquartile range for the number of days between tests, this was defined as the number of days since the previous SHS attendance involving an STI test for that person (this included testing episodes from 2023 and 2024).

We used proxy measures for the eligibility criteria for recommending quarterly testing outlined in national testing guidelines (primarily intended for GBMSM and people using HIV-PrEP) to create an aggregate measure of high STI risk [3,19], more detail in appendix A. This includes HIV-PrEP use, a bacterial STI diagnosis (chlamydia, gonorrhoea or syphilis) in the previous 12 months, and multiple partners (2+) in the previous 3 months. We calculated the number of individuals with markers of high sexual risk by testing frequency category and gender/sexual orientation.

Finally, we calculated the proportion of tests that resulted in a diagnosis of a bacterial STI (positivity). We restricted to bacterial STIs to gain an estimate of diagnoses that would lead to subsequent antibiotic use. We calculated both the positivity of single testing episodes and the proportion of individuals who had at least one positive test in 2024. We examined this by testing frequency category and symptomatic status. As a sensitivity analysis, we assessed whether patterns differed between online and face-to-face settings.

All analysis was completed using STATA v18.0 (StataCorp LP, College Station, TX).

## Results

Overall, there were 2,222,028 episodes in England in 2024 that involved tests for chlamydia, gonorrhoea, syphilis and/or HIV (both online and face-to-face), which constituted 1,750,625 individuals. Symptoms were reported in 18.6% (413,479/2,222,028) of episodes, and 66.8% (1,484,510/2,222,028) were asymptomatic. Partner notification represented 2.6% of testing episodes (58,151/2,222,028).

### Testing combinations

Of the total testing episodes (shown in figure 1 and appendix B), the largest proportion of episodes (66.1% [1,469,194/2,222,028]) were full screens (a chlamydia, gonorrhoea, syphilis and HIV test), followed by episodes with a chlamydia/gonorrhoea test (28.2% [626,747/2,222,028]). When disaggregated by sexual orientation and gender, the group with the largest proportion of full screens were GBMSM (78.7% of testing episodes [297,345/377,945]), followed by heterosexual men (72.5% [365,368/503,754]). Women had the lowest proportion of full screens (60.2% [630,768/1,047,521]) but the highest proportion of dual chlamydia/gonorrhoea tests (36.5% [382,179/1,047,521]). There were differences in testing combinations when comparing testing in online versus face-to-face settings (see figure 1).

### Testing frequency

Most episodes were completed by people attending SHSs once in 2024, with 63.1% of episodes being completed by one-time testers (1,402,285/2,222,028), 30.6% of episodes by people testing 2-3 times (679,798/2,222,028) and 6.3% of episodes being made by people testing 4+ times (139,945/2,222,028). This corresponds to 80.1% of people testing once (1,402,285/1,750,625), 18.0% of people testing 2-3 times (314,698/1,750,625) and 1.9% of people testing 4+ times (frequent testing) (33,642/1,750,625).

When comparing the characteristics of different testing groups (see Table 1), there was a higher proportion of GBMSM testing frequently (6.7% of GBMSM tested 4+ times compared to 1.4% of women and 1.1% of heterosexual men). The youngest age group (15-24 years) had the lowest proportion of frequent testers (1.2%). There was a higher proportion of people testing frequently who lived in London (3.6%), who tested online (2.5%), people using HIV-PrEP (13%), and people with 5+ partners in the previous 3 months (10.6%).

Amongst those reporting markers of higher sexual risk (multiple partners in the last 3 months, use of HIV-PrEP or a bacterial STI diagnosis in the 12 months), we found that 15.3% of heterosexual men, 47.1% of GBMSM and 10.5% of women reported at least one of these behaviours (shown in table 2). Overall, we found that 5.8% of these individuals tested 4+ times. A higher proportion of GBMSM reporting higher-risk behaviours tested 4+ times (10.5%), while 44.7% of this group only tested once. The majority of heterosexual men (79.5%) and women (68.4%) reporting higher-risk behaviours only tested once in 2024, and 3.6% of women and 1.6% of heterosexual men who met these criteria tested at least four times. Among those who did not have markers of higher sexual risk, 4.0% of GBMSM, 1.0% of heterosexual men and 1.2% of women were frequent testers.

There was a median of 151 (IQR: 92-247) days between tests. Among people who tested once in 2024, 20% also tested in 2023, these people had a median of 297 (IQR:192-397) days since their previous test. There was a median of 133 days (IQR:91-195) between tests in those testing 2-3 times and 84 days (IQR:62-105) in those testing 4+ times. Those who only had attendances with STI-related symptoms had the highest median number of days between tests (221 [IQR:125-352]).

**Table 1.** Distribution of the time between successive STI/HIV tests in people accessing sexual health services in England by demographic, clinical and behavioural characteristics, 2024.

|  | Median number of days between tests (interquartile range) * | Testing frequency in 2024 * |  |  |  |
|---|---|---|---|---|---|
|  |  | 1 test | 2-3 tests | 4+ tests | Total |
|  | 151 (92-247) | 1,402,285 (80.1%) | 314,698 (18.0%) | 33,642 (1.9%) | 1,750,625 (100.0%) |
| **Age group (years)** |  |  |  |  |  |
| 15-24 | 150 (91-254) | 428,554 (82.4%) | 85,389 (16.4%) | 6,276 (1.2%) | 520,219 (100.0%) |
| 25-34 | 152 (92-250) | 548,762 (78.8%) | 133,248 (19.1%) | 14,658 (2.1%) | 696,668 (100.0%) |
| 35-44 | 152 (92-245) | 268,213 (79.8%) | 60,500 (18.0%) | 7,394 (2.2%) | 336,107 (100.0%) |
| 45+ | 146 (92-231) | 156,722 (79.3%) | 35,557 (18.0%) | 5,314 (2.7%) | 197,593 (100.0%) |
| **Sexual orientation and gender identity** |  |  |  |  |  |
| Heterosexual Males | 166 (95-275) | 359,844 (85.1%) | 58,489 (13.8%) | 4,516 (1.1%) | 422,849 (100.0%) |
| GBMSM | 124 (87-203) | 137,665 (58.8%) | 80,919 (34.6%) | 15,611 (6.7%) | 234,195 (100.0%) |
| Female | 159 (94-267) | 673,982 (80.7%) | 149,730 (17.9%) | 11,695 (1.4%) | 835,407 (100.0%) |
| Not known | 148 (91-252) | 230,794 (89.4%) | 25,560 (9.9%) | 1,820 (0.7%) | 258,174 (100.0%) |
| **Region of residence** |  |  |  |  |  |
| London | 141 (89-235) | 411,668 (73.1%) | 131,084 (23.3%) | 20,086 (3.6%) | 562,838 (100.0%) |
| West Midlands | 150 (92-254) | 116,259 (81.6%) | 23,995 (16.8%) | 2,260 (1.6%) | 142,514 (100.0%) |
| East Midlands | 161 (98-273) | 98,527 (82.8%) | 19,320 (16.2%) | 1,220 (1.0%) | 119,067 (100.0%) |
| East of England | 166 (105-266) | 118,864 (82.8%) | 23,567 (16.4%) | 1,217 (0.9%) | 143,648 (100.0%) |
| North East | 153 (93-259) | 51,789 (83.5%) | 9,615 (15.5%) | 610 (1.0%) | 62,014 (100.0%) |
| Yorkshire & Humber | 157 (94-269) | 103,338 (81.6%) | 21,696 (17.1%) | 1,594 (1.3%) | 126,628 (100.0%) |
| North West | 151 (94-256) | 158,574 (84.8%) | 26,661 (14.3%) | 1,759 (1.0%) | 186,994 (100.0%) |
| South West | 151 (93-252) | 113,947 (82.5%) | 22,251 (16.1%) | 1,906 (1.4%) | 138,104 (100.0%) |
| South East | 162 (96-263) | 200,946 (85.5%) | 31,573 (13.4%) | 2,595 (1.1%) | 235,114 (100.0%) |
| Unknown | 150 (90-259) | 28,373 (84.2%) | 4,936 (14.7%) | 395 (1.2%) | 33,704 (100.0%) |
| **HIV-PrEP/ HIV status** |  |  |  |  |  |
| Negative/unknown | 153 (92-261) | 1,373,025 (82.4%) | 268,469 (16.1%) | 24,734 (1.5%) | 1,666,228 (100.0%) |
| Using PrEP | 113 (85-181) | 16,268 (26.2%) | 37,814 (60.9%) | 8,063 (13.0%) | 62,145 (100.0%) |
| Person living with HIV (tested positive) | 175 (105-238) | 12,992 (58.4%) | 8,415 (37.8%) | 845 (3.8%) | 22,252 (100.0%) |
| **Symptomatic status during attendances †** |  |  |  |  |  |
| Exclusively symptomatic | 221 (125-352) | 287,235 (93.0%) | 20,810 (6.7%) | 712 (0.2%) | 308,757 (100.0%) |
| Exclusively asymptomatic | 153 (92-260) | 906,902 (82.1%) | 179,016 (16.2%) | 19,266 (1.7%) | 1,105,184 (100.0%) |
| Exclusively unknown (never reported) | 157 (93-245) | 208,148 (84.1%) | 36,092 (14.6%) | 3,255 (1.3%) | 247,495 (100.0%) |
| Mixed symptomatic statuses | 114 (78-178) | - | 78,780 (88.3%) | 10,409 (11.7%) | 89,189 (100.0%) |
| **Test setting** |  |  |  |  |  |
| Face-to-face | 151 (91-254) | 741,559 (83.9%) | 130,099 (14.7%) | 12,004 (1.4%) | 883,662 (100.0%) |
| Online | 151 (92-244) | 660,726 (76.2%) | 184,599 (21.3%) | 21,638 (2.5%) | 866,963 (100.0%) |
| **Number of sex partners‡** |  |  |  |  |  |
| Not stated/known/applicable | 153 (92-251) | 1,088,843 (80.0%) | 245,843 (18.1%) | 25,918 (1.9%) | 1,360,604 (100.0%) |
| 0 | 139 (85-235) | 24,090 (76.4%) | 6,550 (20.8%) | 903 (2.9%) | 31,543 (100.0%) |
| 1 | 174 (99-294) | 203,651 (88.2%) | 26,267 (11.4%) | 1,046 (0.4%) | 230,964 (100.0%) |
| 2 to 4 | 126 (85-215) | 71,353 (72.4%) | 24,443 (24.8%) | 2,707 (2.8%) | 98,503 (100.0%) |
| 5 or more | 105 (84-167) | 14,348 (49.5%) | 11,595 (40.0%) | 3,068 (10.6%) | 29,011 (100.0%) |
| **STI positive result in 2024** |  |  |  |  |  |
| No | 158 (95-263) | 1,302,789 (82.2%) | 258,453 (16.3%) | 23,279 (1.5%) | 1,584,521 (100.0%) |
| Yes | 111 (75-182) | 99,496 (59.9%) | 56,245 (33.9%) | 10,363 (6.2%) | 166,104 (100.0%) |
| **STI test due to partner notification** |  |  |  |  |  |
| No | 152 (92-251) | 1,364,654 (80.5%) | 299,138 (17.7%) | 30,508 (1.8%) | 1,694,300 (100.0%) |
| Yes | 113 (76-187) | 37,631 (66.8%) | 15,560 (27.6%) | 3,134 (5.6%) | 56,325 (100.0%) |

\* Categorised number of tests completed in 2024 † Where someone has attended a SHS multiple times but had a different symptomatic status (any combination of symptomatic/asymptomatic/unknown), they have been labelled as having a mixed symptomatic status. ‡ in the previous 3 months

**Table 2.**
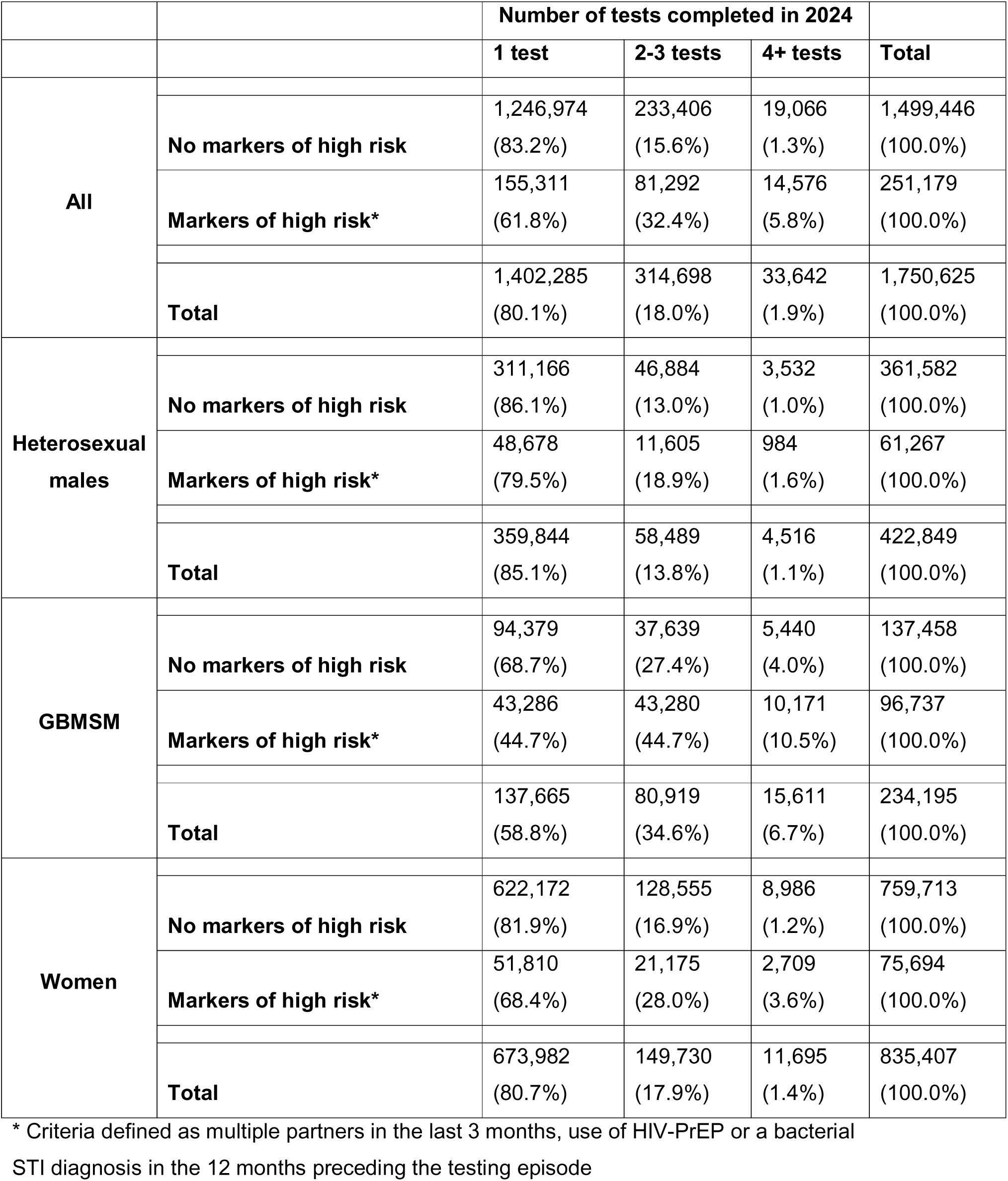
Number of tests completed in 2024 among those with markers of higher STI transmission risk.

### Positivity

Overall, 7.9% of testing episodes in 2024 were positive (had a chlamydia, gonorrhoea or syphilis diagnosis). People who tested once had the lowest proportion of positive diagnoses (7.1% of episodes) and 30.8% of people who tested 4+ times had at least one positive STI diagnosis in 2024. Tests occurring in frequent testers had a higher proportion of positive results (7.1% vs 9.9% [1 test vs 4+ tests]), this difference was higher in tests in people with STI-related symptoms (8.4% vs 13.5%) or an unknown symptomatic status (9.3% vs 14.4%) (figure 2a). There was not a large difference in the proportion of individuals with a positive result when comparing consistently asymptomatic and symptomatic groups (see figure 2b). The groups with the highest proportion of positive results were frequent testers with consistently unreported (37.1%) or mixed symptomatic statuses (35.8%).

When we compared face-to-face and online services, the positivity in face-to-face SHSs was higher overall (9.86% face-to-face vs 6.17% online). However, the patterns described above remained consistent in both settings (appendix C/D)

## Discussion

Our analysis of national STI surveillance data in England found that 80% of individuals accessing SHS in 2024 tested once. Among those who reported markers of high risk of STIs, only 6% were testing frequently (4+ times). This varied by gender identity and sexual orientation, with a higher proportion of GBMSM (10.5%) than women (4%) and heterosexual men (2%) testing frequently. HIV-PrEP users are more likely to be frequent testers, reflecting clinical guidelines for HIV and STI screening[19]. People with more partners also had a higher proportion of frequent testing. This indicates that groups at greater risk of STIs[1, 7] could have higher engagement with STI testing. While a relatively low proportion of those who may be recommended to test more frequently had at least 4 testing episodes in 2024, GBMSM and people using HIV-PrEP often tested at least twice a year (55% and 74% respectively). Among frequent testers, the median interval between testing episodes was 84 days which indicates that testing intervals in this group were, on average, approximately quarterly (every 3 months). However, classification as a frequent tester was based on the number of testing episodes during 2024 and does not demonstrate that an individual tested at regular quarterly intervals.

### Contextualising findings

GBMSM in the UK are recommended to test quarterly if they are at a higher risk of STI transmission. Only a relatively small proportion of GBMSM who reported high-risk behaviours (roughly equivalent to quarterly testing criteria) tested 4+ times in 2024, which could be an indication of unmet need[20]. In a community sample of GBMSM in the UK, 24% of GBMSM who met quarterly testing criteria self-reported that they tested at least 4 times[21]. This is higher than the 10.5% that we report, which could be due to that survey’s use of a convenience sample of GBMSM who were at higher risk of STIs and more likely to engage with SHSs for STI testing.

We found that 13% of HIV-PrEP users tested frequently, with 74% testing at least twice a year. Usage of HIV-PrEP is higher among GBMSM than other groups in England[22] and is recommended among those reporting higher-risk sexual behaviours[19]. Quarterly testing is recommended for HIV-PrEP users[3,19]. Higher rates of testing among GBMSM who were HIV-PrEP users (compared to other GBMSM) have been shown previously[23]. Literature comparing testing frequency among HIV-PrEP users is limited; however, this number is lower than estimates using behavioural surveys in Australia (which also recommends quarterly testing among HIV-PrEP users[23]). It is also possible that a subset of people using HIV-PrEP in our analysis were not in the frequent testing category if they initiated HIV-PrEP use later in the year, as they had reduced opportunity for subsequent tests.

The alignment of more frequent testing with higher behavioural risk was potentially observed in the higher test positivity in frequent testers. However, the larger number of tests likely contributes to this, as they had more opportunities to receive a positive result. More frequent testing among asymptomatic people provides more opportunity for diagnoses (and subsequent treatment) of asymptomatic infections that may have no resulting morbidity. Despite this, it also appears frequent testers display a higher level of sexual risk, so this group may have a higher prevalence of STIs. Quarterly asymptomatic testing may also be recommended for the year following an STI diagnosis[2], which may increase the number of tests in those with a positive result.

There were also a minority who tested frequently despite not reporting behavioural markers equivalent to quarterly testing criteria (4% of GBMSM without recorded risk factors); however, these people may have reported high-risk behaviours to a clinician but were not recorded as such in GUMCAD. When we applied the same proxy quarterly testing eligibility criteria to women and heterosexual men we found that a small proportion of those meeting the criteria were also testing frequently (4% of women and 2% of heterosexual men). This indicates a possible self-identification of risk resulting in frequent STI testing, although this is occurring less frequently than among GBMSM.

The recommended minimum investigations, for all people attending SHSs for STI related needs, even if asymptomatic, are tests for chlamydia, gonorrhoea, syphilis and HIV, and most testing episodes recorded full screening. A substantial portion (28%) of tests were dual chlamydia/gonorrhoea only tests, reflecting common use of dual nucleic acid amplification tests (NAATs) on the same sample. The higher proportion of these tests through online testing compared to face-to-face, will partly be due to a high proportion of chlamydia screening being accessed online[7], but it also may reflect difficulty returning the blood samples required for online postal self-service syphilis and HIV testing[6,18] or these people being asymptomatic or displaying lower risk.

There were also a lower proportion of young people in the frequent testing group. While young people have disproportionately high STI diagnosis rates, recent patterns in England show a reduced level of STI testing in young people following the COVID-19 pandemic[24]. Our findings indicate that there are a large number of young people who tested for bacterial STIs, but they may only be testing annually. While there is a generally higher level of STI diagnosis in young people[7], annual testing will be suitable for a large proportion of young people who display lower levels of risk.

Generally, there is a higher rate of STI testing in London compared to the rest of England[24]. Residents of London also had a higher proportion of frequent testers than other regions. This possibly represents lower barriers to SHS access in London, such as a relatively higher density of SHS, the transport infrastructure, and long-established city-wide online postal self-service commissioning[20]. This could also be a result of the higher population of GBMSM in London[25], who represent a larger proportion of the frequent testing group.

### Strengths and Limitations

The use of national surveillance data provides a comprehensive description of testing behaviours in people accessing SHSs in England. However, there are limitations associated with the use of GUMCAD data. We cannot trace individuals between clinics, so we have likely under-estimated testing frequency if individuals have used multiple SHSs. While we cannot quantify the people who utilise multiple SHSs, combining the surveillance data presented here with behavioural surveys, such as those described above[21] provides plausible datasets for triangulation of findings. Some people may have also used STI testing outside SHSs (such as in general practices or private providers), which are not captured within GUMCAD. Additionally, restricting analyses to a single calendar year may have limited our ability to accurately identify frequent testers who tested multiple times across calendar years.

The level of data completion within GUMCAD is generally high; however, some variables (symptomatic status and number of partners) introduced more recently have a comparatively higher number of missing records[18, 26]. Thus, there may be individuals meeting quarterly testing criteria that were not identified. The proportion of SHSs reporting these newer variables is also lower in London. These SHSs report a high number of bacterial STI diagnoses, which will contribute to the high test-positivity in attendances with unknown symptomatic status. We will be able to report this more accurately in the future as data quality improves[18, 26].

### Policy implications

We found that only a minority of those meeting criteria for quarterly testing had four or more recorded testing episodes during 2024. Some countries are reassessing asymptomatic screening guidelines for chlamydia and gonorrhoea, particularly in response to AMR[8, 10, 13]. There have been mixed responses from GBMSM and professional stakeholders to a potential reduction in asymptomatic screening[8]. It is important that any changes to asymptomatic STI screening guidance in GBMSM should involve stakeholders and acknowledge that actual testing frequency is lower than recommended in some instances (37.0% of GBMSM accessing SHS in 2024 appear to be testing less frequently than they would be recommended, and only 2.3% appear to be testing more frequently than they may be recommended), so reducing the recommended frequency may have limited service impact.

Another aspect that should be considered is the potentially different testing needs of men who have sex with both men and women[11]. This is due to the potential impact of adverse events related to chlamydia and gonorrhoea in women[3]. To reduce the impact of these infections, it may be important to empower women and men who have sex with women to identify any high-risk behaviours and to mitigate this by testing for STIs when necessary.

## Conclusion

Overall, a majority of individuals accessing SHS in England only had one STI test in 2024. While frequent testing was more common among people with increased risk of STIs, only a minority of people with these markers tested 4+ times in 2024. Gaining an insight into testing frequency and motivation is important to inform priority groups for interventions and messaging focused on STI testing. As STI testing guidelines in the UK evolve, it remains critical to use the best available surveillance and research evidence, with community and clinical stakeholder input, to minimise the harms associated with STIs.

## Acknowledgement

We would like to offer our thanks to all the sexual health services that report GUMCAD data to UKHSA.

## Data availability statement

All data were collected within statutory approvals granted to the UK Health Security Agency for infectious disease surveillance and control. The UK Health Security Agency has approval to handle data obtained by the GUMCAD STI surveillance system under Regulation 3 of the Health Service (Control of Patient Information) Regulations 2002. Information was held securely and in accordance with the Data Protection Act 2018 and Caldicott guidelines.

## Competing interests

None to declare

## Funding

George Baldry receives funding from the Medical Research Council (grant number: MR/W006774/1). The funders had no role in study design, data collection and analysis, decision to publish, or preparation of the manuscript. All other authors received no specific funding for this work.

## Author contributions

GB, AKH, HM, SJM and KS curated an analysis plan with support from LF, DO, HF and JS. GB carried out the analysis and write-up of the first manuscript draft with support from AKH. All authors contributed to successive drafts and reviewed and approved the final manuscript.

## Ethical approval

In its role providing infectious disease surveillance, UK Health Security Agency has permission to handle data obtained through the GUMCAD STI Surveillance System under Regulation 3 of the Health Service (Control of Patient Information) Regulations 2002.

Requests for aggregate data can be made by contacting the GUMCAD, where all publicly released data must adhere to UKHSA data sharing guidelines.

This analysis has been subject to an internal review which considered the study design, content, and feasibility. The review also covered all legal, financial, regulatory, and ethical considerations. As a result of this review, the study was categorized as surveillance and as no ethical issues were identified it was decided that review by an ethics committee would not be necessary.

## Notes

### Competing Interest Statement

The authors have declared no competing interest.

### Author Declarations

In its role providing infectious disease surveillance, UK Health Security Agency has permission to handle data obtained through the GUMCAD STI Surveillance System under Regulation 3 of the Health Service (Control of Patient Information) Regulations 2002. This analysis has been subject to an internal review which considered the study design, content, and feasibility. The review also covered all legal, financial, regulatory, and ethical considerations. As a result of this review, the study was categorized as surveillance and as no ethical issues were identified it was decided that review by an ethics committee would not be necessary.

### Summary of Updates

Manuscript text updated, new appendix added, abstract edited

