## Appendix A for "Frequency of bacterial STI testing amongst people accessing sexual health services in England, 2024: a cross-sectional analysis of national surveillance data"

The British Association of Sexual Health and HIV (BASHH) national guidelines recommend annual STI and HIV testing for sexually active GBMSM, with three-monthly testing recommended for those reporting higher-risk sexual behaviours. This includes multiple or anonymous sexual partners in the previous 12 months, condomless anal sex with partners of unknown or serodiscordant HIV status in the previous 12 months, and drug use during sex (chemsex) in the past 6 months[1]. Regular STI testing is also recommended for people using HIV-PrEP who have new or multiple sex partners[2].

These recommendations informed the selection of proxy measures available in GUMCAD to create an aggregate measure of high STI risk. The measures included two or more sexual partners in the previous three months, a bacterial STI diagnosis (chlamydia, gonorrhoea or syphilis) in the previous 12 months, and current HIV-PrEP use. Previous bacterial STI diagnosis was used as a marker of increased STI risk, rather than to establish that all individuals with a previous diagnosis should be independently recommended quarterly bacterial STI testing. Although the behavioural testing-frequency recommendations were primarily developed for GBMSM, these proxy measures were applied across all SHS attendees in the

analysis.
